# Interplay of IL6 and CRIM1 on thiopurine-induced neutropenia in leukemic patients with wild-type NUDT15 and TPMT

**DOI:** 10.1101/2020.07.21.20158931

**Authors:** Hyery Kim, Seungwon You, Yoomi Park, Jung Yoon Choi, Youngeun Ma, Kyung Tak Hong, Kyung-Nam Koh, Sunmin Yun, Kye Hwa Lee, Hee Young Shin, Suehyun Lee, Keon Hee Yoo, Ho Joon Im, Hyoung Jin Kang, Ju Han Kim

**Affiliations:** Department of Pediatrics, Asan Medical Center, University of Ulsan College of Medicine, Seoul, Korea; Seoul National University Biomedical Informatics (SNUBI), Division of Biomedical Informatics, Seoul National University College of Medicine, Seoul 03080, Korea; Department of Pediatrics, Seoul National University College of Medicine, Seoul 03080, Korea; Seoul National University Cancer Research Institute, Seoul, Korea; Department of Pediatrics, Seoul National University Bundang Hospital, Seoul, Korea; Department of Information Medicine, Asan Medical Center and University of Ulsan College of Medicine, Seoul 05505, Korea; Department of Biomedical Informatics, College of Medicine, Konyang University, Daejeon, Korea; Department of Pediatrics, Samsung Medical Center, Sungkyunkwan University School of Medicine, Seoul, Korea

**Keywords:** IL6, CRIM1, thiopurine toxicity, genetic determinant

## Abstract

**Background:** *NUDT15* and *TPMT* variants are strong genetic determinants of thiopurine-induced hematological toxicity. Despite recent discovery of homozygous *CRIM1* effect on thiopurine toxicity, many patients with wild-type *NUDT15, TPMT*, and *CRIM1* still suffer from thiopurine toxicity, and therapeutic failure and relapse of acute lymphoblastic leukemia (ALL).

**Methods:** Novel PGx interactions associated with thiopurine toxicity in 320 pediatric ALL patients were investigated using whole-exome sequencing technology for the last-cycle 6-Mercaptopurine dose intensity percentage (DIP) tolerated by pediatric ALL patients.

**Results:** *IL6* rs13306435 carriers (*N*=19) exhibited significantly lower DIP (48.0±27.3%) than non-carriers (*N*=209, 69.9±29.0%; *p*=0.0016 and 0.0028 by *t*-test and multiple linear regression, respectively). Of the 19 carriers, seven with both heterozygous *IL6* rs13306435 and *CRIM1* rs3821169 showed significantly decreased DIP (24.7±8.9%) than those with *IL6* (*N*=12, 61.6±25.1%) or *CRIM1* (*N*=94, 68.1±28.4%) variant only. Both *IL6* and *CRIM1* variants showed marked inter-ethnic variability. Significant interplay between *IL6* and *CRIM1* in thiopurine toxicity was suggested. GVB (Gene-wise Variant Burden)-based four-gene-interplay model showed the best odds ratio (8.06) and potential population impact (i.e., relative risk (5.73), population attributable fraction (58%), number needed to treat (3.67) and number needed to genotype (12.50)).

**Conclusions:** Interplay of *IL6* rs13306435 and *CRIM1* rs3821169 was suggested as independent and/or additive genetic determinant of thiopurine toxicity beyond *NUDT15* and *TPMT* in pediatric ALL.

## Introduction

Despite improvements of combination drug therapy and risk stratification, about 20% of pediatric acute lymphoblastic leukemia (ALL) patients still suffer from drug resistance and treatment failure due to drug toxicities. In European populations, about 50% of thiopurine-induced cytotoxic adverse reactions such as severe neutropenia and leukopenia are explained by *NUDT15* and *TPMT* genetic variants [1, 2, 3, 4]. The Clinical Pharmacogenetics Implementation Consortium (CPIC) [5] publishes practical guidelines for the implementation of pharmacogenetic (PGx) testing of thiopurine by using traditional star (*) allele-based molecular phenotyping for *NUDT15* and *TPMT* [6,7].

According to the established guideline, thiopurine dose is pharmacogenetically titrated based on the known risk variants of *NUDT15* and *TPMT*. However, a substantial proportion of leukemia patients who have no genetic variation in *NUDT15* or *TPMT* still suffer from life-threatening toxicity, which may result in dose reduction and/or discontinuation of thiopurine, resulting therapeutic failure and relapse of leukemia. In an attempt to overcome the PGx gap, *CRIM1* rs3821169 homozygote in East Asians has been reported as a novel risk variant of thiopurine toxicity [8]. Heterozygotes of the variant showed only mild effect on thiopurine toxicity with unknown clinical impact. However, its high prevalence (T=0.066, the Phase 3 of the 1000 Genomes Project [9]) and remarkable inter-ethnic variability (see Table 2) might have severely confounded previous PGx studies for thiopurine toxicity. Therefore, investigating PGx interactions of novel genes/variants other than *NUDT15* and *TPMT* variations is urgently needed for preventing thiopurine toxicity and improving pediatric ALL care.

**Table 2.**
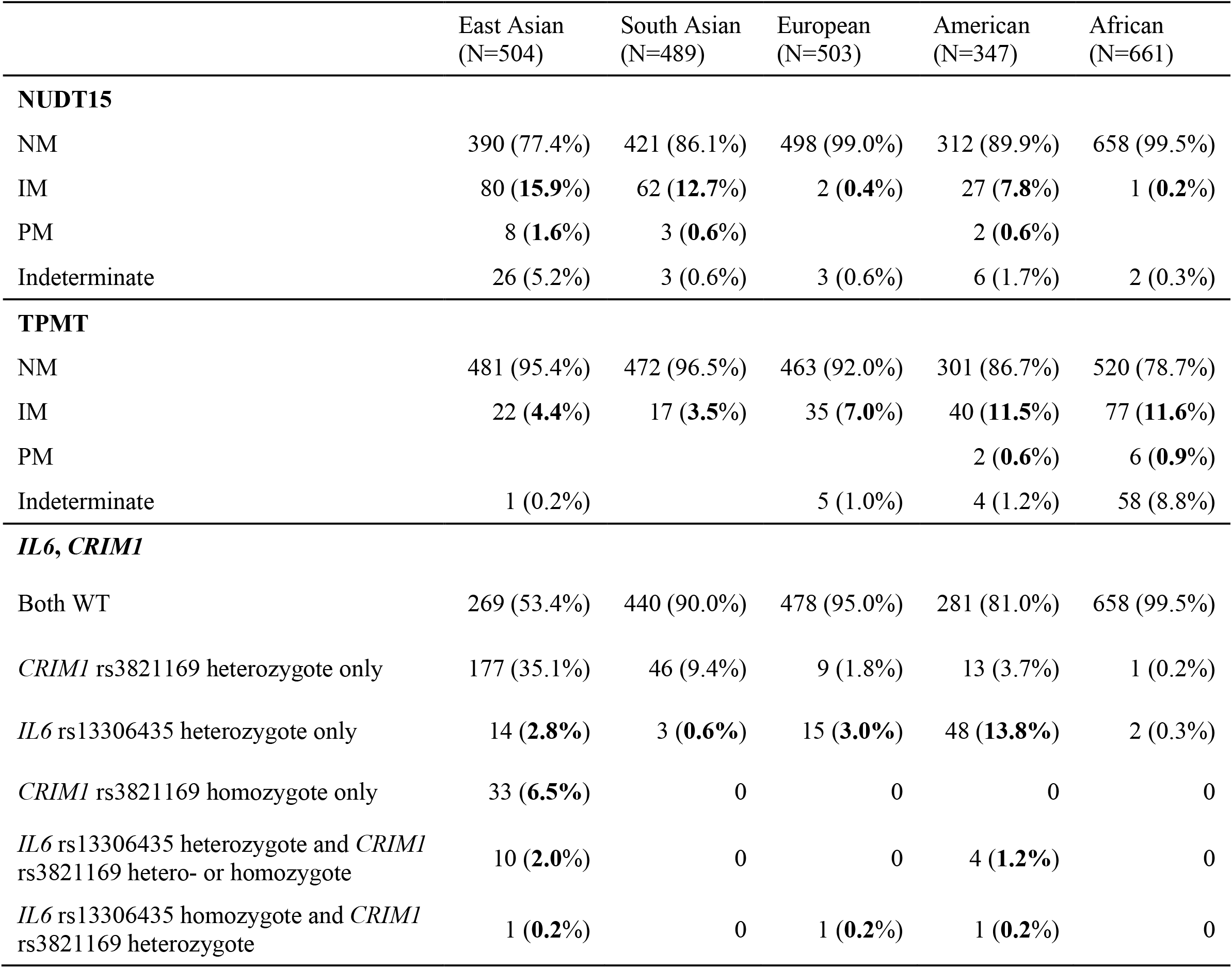
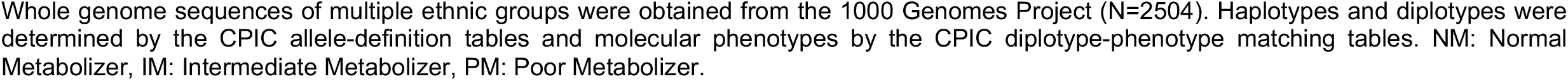
Inter-ethnic variability of thiopurine toxicity-associated pharmacogenetic variants.

|  | East Asian<br>(N=504) | South Asian<br>(N=489) | European<br>(N=503) | American<br>(N=347) | African<br>(N=661) |
| --- | --- | --- | --- | --- | --- |
| **NUDT15** |  |  |  |  |  |
| NM | 390 (77.4%) | 421 (86.1%) | 498 (99.0%) | 312 (89.9%) | 658 (99.5%) |
| IM | 80 (**15.9%**) | 62 (**12.7%**) | 2 (**0.4%**) | 27 (**7.8%**) | 1 (**0.2%**) |
| PM | 8 (**1.6%**) | 3 (**0.6%**) |  | 2 (**0.6%**) |  |
| Indeterminate | 26 (5.2%) | 3 (0.6%) | 3 (0.6%) | 6 (1.7%) | 2 (0.3%) |
| **TPMT** |  |  |  |  |  |
| NM | 481 (95.4%) | 472 (96.5%) | 463 (92.0%) | 301 (86.7%) | 520 (78.7%) |
| IM | 22 (**4.4%**) | 17 (**3.5%**) | 35 (**7.0%**) | 40 (**11.5%**) | 77 (**11.6%**) |
| PM |  |  |  | 2 (**0.6%**) | 6 (**0.9%**) |
| Indeterminate | 1 (0.2%) |  | 5 (1.0%) | 4 (1.2%) | 58 (8.8%) |
| ***IL6, CRIM1*** |  |  |  |  |  |
| Both WT | 269 (53.4%) | 440 (90.0%) | 478 (95.0%) | 281 (81.0%) | 658 (99.5%) |
| *CRIM1* rs3821169 heterozygote only | 177 (35.1%) | 46 (9.4%) | 9 (1.8%) | 13 (3.7%) | 1 (0.2%) |
| *IL6* rs13306435 heterozygote only | 14 (**2.8%**) | 3 (**0.6%**) | 15 (**3.0%**) | 48 (**13.8%**) | 2 (0.3%) |
| *CRIM1* rs3821169 homozygote only | 33 (**6.5%**) | 0 | 0 | 0 | 0 |
| *IL6* rs13306435 heterozygote and *CRIM1* rs3821169 hetero- or homozygote | 10 (**2.0%**) | 0 | 0 | 4 (**1.2%**) | 0 |
| *IL6* rs13306435 homozygote and *CRIM1* rs3821169 heterozygote | 1 (**0.2%**) | 0 | 1 (**0.2%**) | 1 (**0.2%**) | 0 |

Whole genome sequences of multiple ethnic groups were obtained from the 1000 Genomes Project (N=2504). Haplotypes and diplotypes were determined by the CPIC allele-definition tables and molecular phenotypes by the CPIC diplotype-phenotype matching tables. NM: Normal Metabolizer, IM: Intermediate Metabolizer, PM: Poor Metabolizer.

The categorical nature of the traditional star allele haplotype-based method can be complemented by the quantitative nature of Gene-wise Variant Burden (GVB) method for evaluating complex interplays of multiple genes/variants [10]. For instance, designating three categories (i.e., poor (PM), intermediate (IM), and normal (NM) metabolizers) per a gene creates exponentially increasing complexity of 3^*N*^ for a drug with *N*-gene PGx interactions. *NUDT15* and *TPMT* have already required nine PGx subgroups for thiopurine, which will increase exponentially by new PGx discoveries across different ethnic groups. GVB quantitates the cumulative variant burden of one or more genes into a single score with dimensionality reduction and hence to provide a reliable frame for multiple gene-interaction analysis [11, 12, 13].

The present study aimed to identify novel PGx interactions associated with thiopurine toxicity in pediatric ALL patients who carry both wild-type (WT) *NUDT15* and *TPMT* (and do not carry *CRIM1* rs3821169 homozygote) by using whole-exome sequencing (WES) technology. We identified and evaluated the deterministic effect and their interaction of novel candidate PGx variants on a clinically important hematological toxicity indicator: the last-cycle 6-MP dose intensity percentage (DIP) tolerated by pediatric ALL patients. We provided not only the measures of clinical validity but also the measures of population impact (or clinical utility) including relative risk (RR), population attributable fraction (PAF), number needed to treat (NNT), and number needed to genotype (NNG) [14] for preventing thiopurine toxicity.

## Materials and Methods

### Subjects

We recruited 320 Korean pediatric ALL patients who underwent maintenance therapy with 6-Mercaptopurine (6-MP) from three teaching hospitals, Seoul National University Hospital (SNUH), Asan Medical Center (AMC), and Samsung Seoul Medical Center (SMC), located in Seoul, South Korea. All of the subjects conformed with the exclusion criteria (i.e., relapse of the disease, stem cell transplantation, Burkitt’s lymphoma, mixed phenotype acute leukemia, infant ALL, or very high risk of ALL). Patients allocated to standard risk group were treated with Children’s Cancer Group (CCG)-1891 [15], CCG-1952 [16] or Children’s Oncology Group AALL-0331 regimens [17]. In high risk group, CCG-1882 [18], 0601 or 1501 protocols for Korean multicenter studies [19] were used. In Korea, the planned dose of 6-MP was modified from 75 to 50 mg/m^2^, as many patients who had been given the same dose under the original Western protocol exhibited moderate to severe toxicities during 6-MP administration [20, 21]. Doses of 6-MP during maintenance were adjusted to maintain a WBC count of 2.0-3.5×10^9^/L with an absolute neutrophil count (ANC) over 500/μL, and hepatotoxicity related dose modifications were performed at the discretion of the treating physician. Hematological toxicity as the clinical endpoint was estimated by the tolerated last-cycle 6-MP DIP (%). The percentage of the actually prescribed amount to the planned dose was defined as the last cycle 6-MP DIP by using the recorded 6-MP dose per meter body surface area over the last cycle (12-week) of maintenance. The doses of the last maintenance cycle were considered because dose modification of 6-MP was mainly adopted during early phase of maintenance. Further detailed description of patients and the measurements are summarized in our previous study [8, 20, 21]. The present study was approved by the SNUH, AMC, and SMC Institutional Review Boards. Written informed consent was obtained from each participant.

### Whole-exome sequencing and pharmacogenomic subgrouping

WES data were obtained from pediatric ALL patients and analyzed in a bioinformatics pipeline as previously described [8, 10, 11]. CPIC provides major PGx genes with haplotype definitions and molecular function annotations based on star (*) nomenclature. We classified ALL patients into PM, IM, and NM groups of *NUDT15* and *TPMT* according to CPIC classifications [6, 7]. We considered NMs as WTs for both genes. We found that no star name designated yet to novel (or candidate) PGx genes. Thus, for the purpose of the present study, we defined WTs for *CRIM1* and *IL6* as non-carriers of *CRIM1* rs3821169 homozygote and *IL6* rs13306435 hetero/homozygote, respectively (Table 1). Haplotypes were determined by PHASE 2.1.1 [22, 23].

**Table 1.**
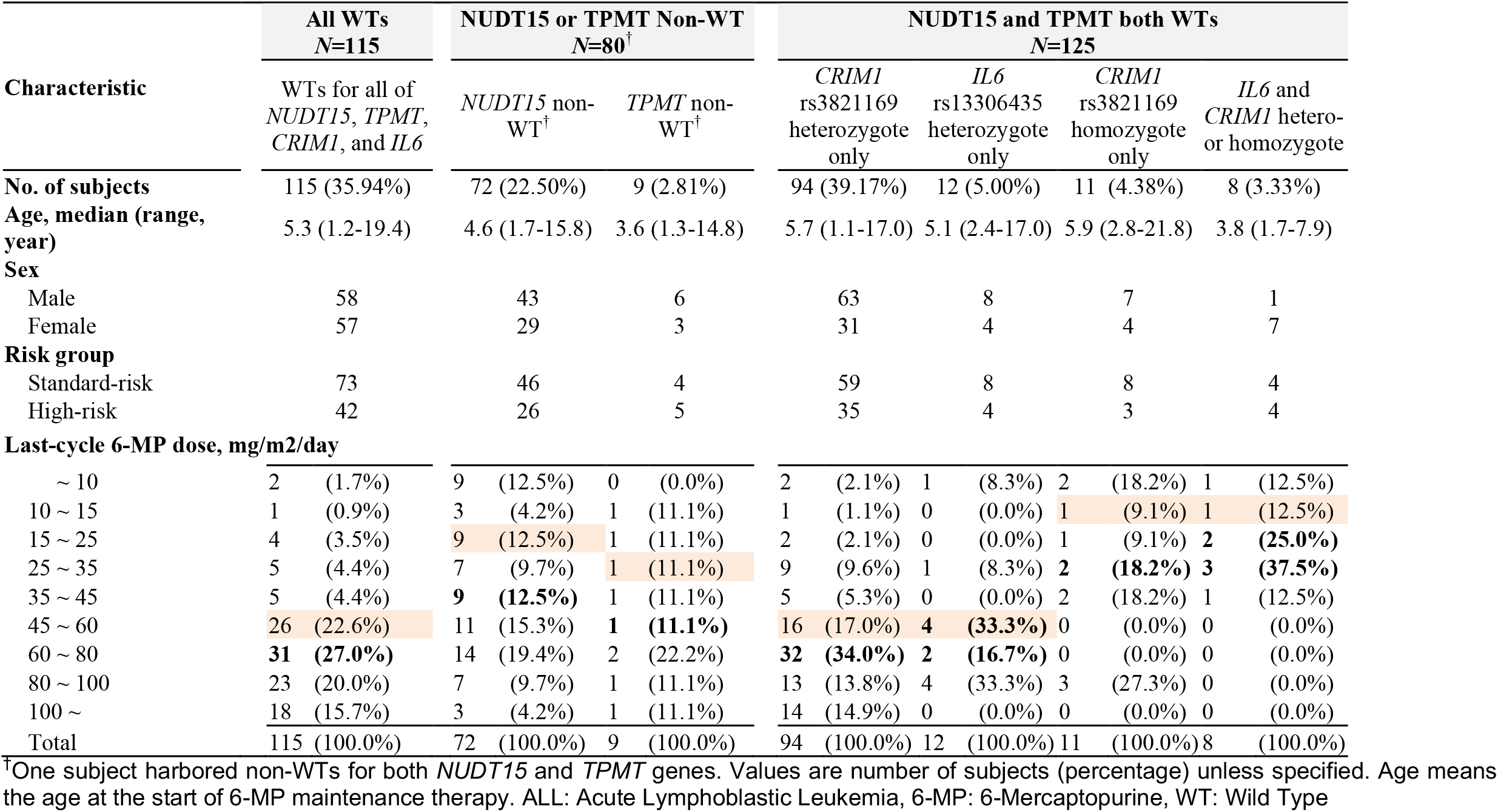
Clinical characteristics of 320 pediatric ALL patients with 6-MP maintenance therapy according to their pharmacogenetic subgroups of *NUDT15, TPMT, CRIM1* and IL6 genes.

### Gene-wise variant burden for evaluating single- and multi-gene effects

Gene-wise variant burden (GVB) analysis was performed to evaluate the aggregated impact of both common and rare variants [10, 11]. The GVB of a coding gene for each individual was defined as the geometric mean of the SIFT (Sorting Intolerant From Tolerant) [24] scores of the coding variants (with SIFT score <0.7) in the coding gene, where GVB^*G*^ denotes the GVB score of gene *G* [range: 0.0∼1.0]. The more deleterious variant burden, the lower the score. Multigene effect was evaluated by defining GVB^*A,B,C*^ as the geometric mean of GVB^*A*^, GVB^*B*^ and GVB^*C*^ [range: 0.0∼1.0]. Gene-variant interaction was considered by defining conditional GVB^*G^*(*variant*)^ as the GVB score of gene *G* dependent on the presence or absence of the specified *variant*. For example, GVB^*CRIM1^ (rs13306435)*^ equals GVB^*CRIM1*^ when *rs13306435* is present and vanishes to a WT score of 1.0 when absent.

### Inter-ethnic variability of allele frequencies and molecular phenotypes

By using the 2504 whole genome sequences with multiple ethnicities provided by the 1000 Genomes Project phase 3 [9], we investigated inter-ethnic distributions of the PGx alleles and haplotypes with their molecular phenotypes associated with thiopurine toxicity (see Table 2).

### Statistical analysis

The last-cycle 6-MP DIPs (%) according to different PGx groups were tested by Student’s *t*-test or one-way ANOVA with posthoc Tukey test. Multiple linear regression was also applied to adjust confounding clinical variables. The powers of GVB^*NUDT15*^, GVB^*TPMT*^, GVB^*CRIM1*^, and GVB^*IL6*^, and their combinations for predicting 6-MP DIP were systematically evaluated by analyzing ROC (receiver operating characteristic) curves across eight different DIP cutoffs (i.e., 10%, 15%, 25%, 35%, 45%, 60%, 80%, and 100%) in terms of AUCs (areas under the ROC curves) (see Figs. 3 and 4). An ROC curve is a two-dimensional depiction of classification performance integrating all sensitivity and specificity values at all cutoff levels [25]. All statistical analyses were performed using the R statistical package (version 3.5.1). R package ‘pROC’ was used for calculating AUC values [26]. The optimal cutoff for the GVB score was determined by maximizing Youden’s index [27]

**Figure 3.**
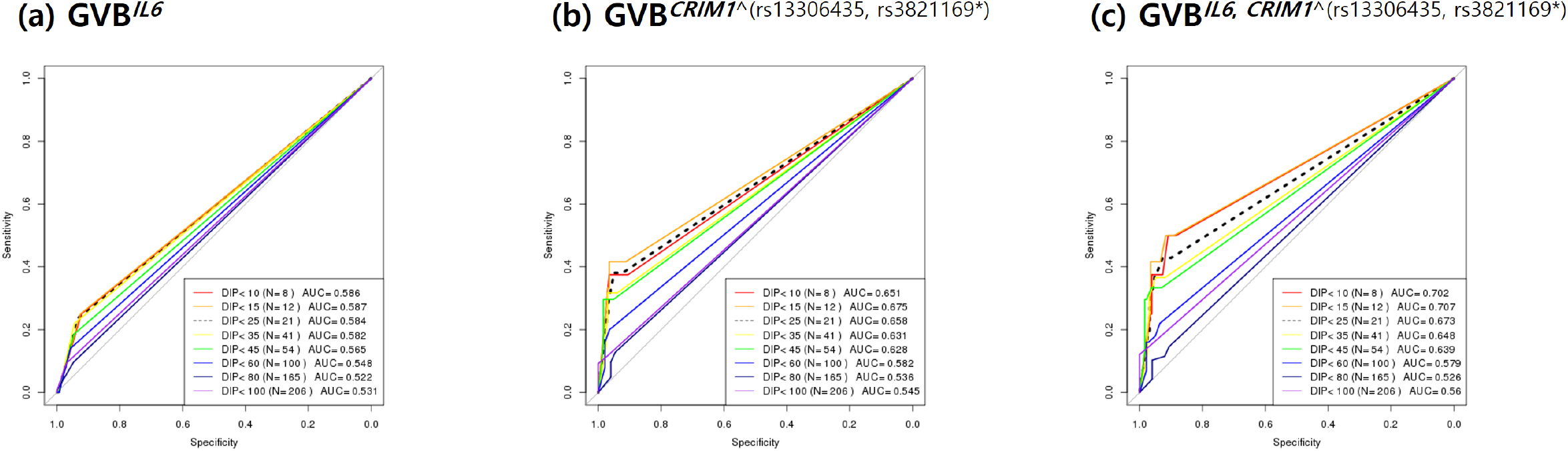
ROC analysis of *IL6, CRIM1*, and their combined prediction accuracies for thiopurine intolerance among pediatric ALL subjects with both wild-type *NUDT15* and *TPMT* genes (N=240). Single-gene prediction models of (a) *IL6* and (b) *CRIM1* were highly outperformed by (c) the two-gene combined model for predicting thiopurine intolerance at all DIP levels. We excluded 80 among the total of 320 subjects to control the effects of the well-established *NUDT15* and *TPMT* genes and performed ROC analysis for the 240 subjects with both wild-type *NUDT15* or *TPMT* genes. Prediction accuracies were measured across 8 cutoff levels for the last-cycle 6-MP DIP (%) (≤10%, ≤15%, ≤25%, ≤35%, ≤45%, ≤60%, ≤80%, and ≤100%) in terms of AUC. DIP (%): Dose Intensity Percentage, AUC: Area Under the ROC (Receiver Operating Characteristics) Curve, GVB: Gene-wise Variant Burden, GVB^*CRIM1^(rs13306435, rs3821169*)*^: GVB of *CRIM1* dependent on *IL6* rs13306435 or *CRIM1* rs3821169 homozygote.

**Figure 4.**
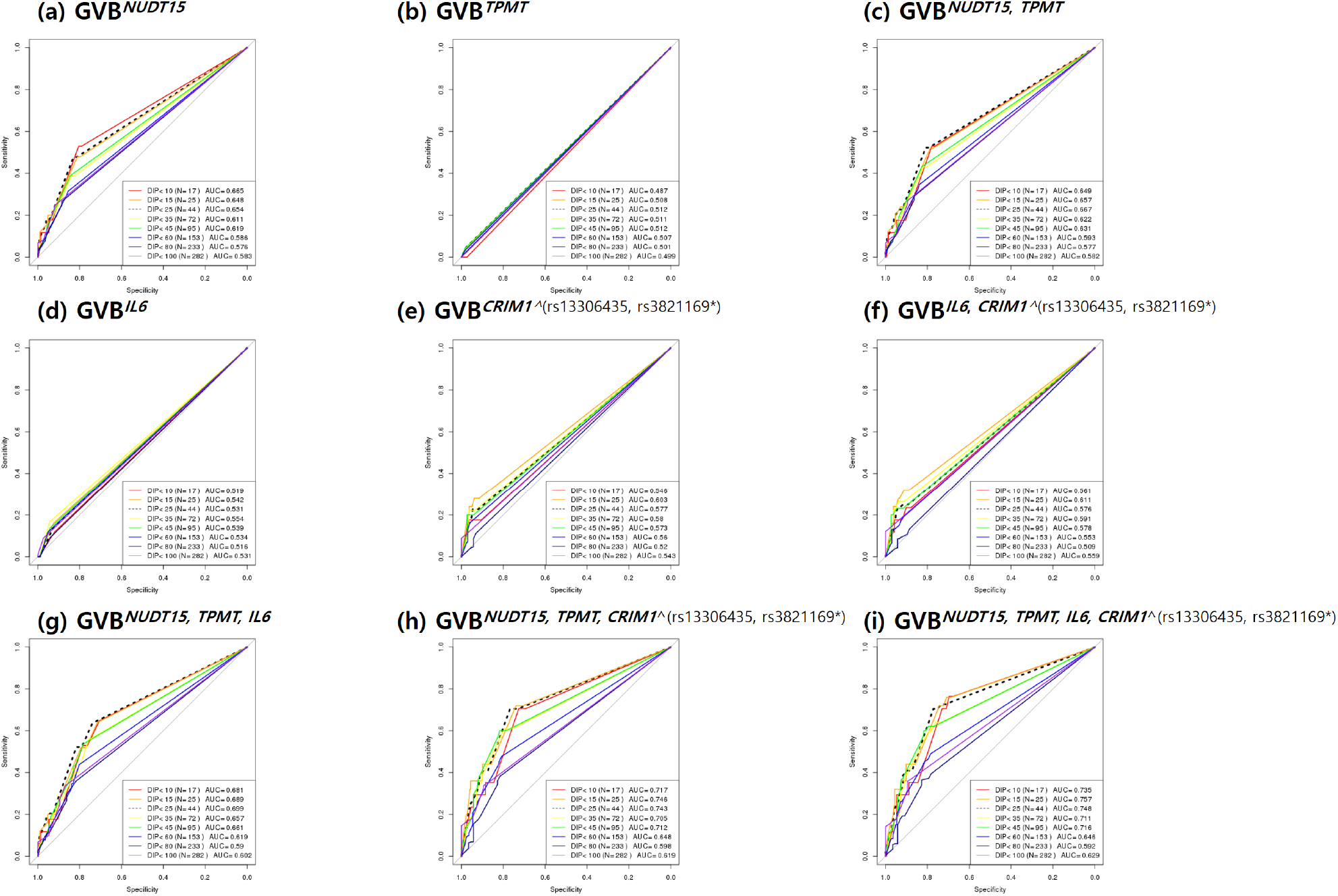
Prediction accuracy profile of single- and multi-gene models for thiopurine intolerance in the whole pediatric ALL subjects (N=320). Single-gene prediction models of (a) NUDT15 and (b) TPMT were outperformed by (c) the two-gene combined model and of (d) IL6 and (e) CRIM by (f) IL6-CRIM1 combined model. Three-gene models of NUDT15, TPMT and (g) IL6 and (h) CRIM1 were outperformed by (i) all four-gene combined model. Overall the final (i) four-gene combined model outperformed other models for predicting thiopurine intolerance at all DIP levels in pediatric ALL subjects (N=320). AUCs for predicting the last-cycle 6-MP DIP (%) were measured at 8 cutoff levels of (≤10%, ≤15%, ≤25%, ≤35%, ≤45%, ≤60%, ≤80%, and ≤100%). ALL: Acute Lymphoblastic Leukemia, DIP: Dose Intensity Percentage, AUC: Area Under the Receiver Operating Characteristics Curve, GVB: Gene-wise Variant Burden.

GVB^*CRIM1^*(*rs13306435*\*)^ was applied to control the potential confounding effect of the impressively high carrier frequency in East Asians (43.7% (=220/504)) compared to other ethnicities (0.2∼9.4%) and of mild effect of heterozygote on thiopurine toxicity. GVB^*CRIM1^*(*rs13306435*\*)^ denotes a conditional GVB score of *CRIM1* dependent on the presence or absence of homozygous *rs3821169* variant (denoted as *rs3821169\**). It equals GVB^*CRIM1*^ when the subject carries homozygous *rs3821169* variant and otherwise vanishes to 1.0.

## Results

### *IL6* rs13306435 as a novel pharmacogenetic variant on thiopurine toxicity

Table 1 describes clinical characteristics of the whole 320 pediatric ALL patients according to their PGx subgroups; 80 non-WTs (i.e., IMs or PMs) for *NUDT15* (*N*=72) and/or *TPMT* (*N*=9), 115 all WTs (for all of the four genes), and 125 both WTs (for *NUDT15* and *TPMT*) who carried *CRIM1* rs3821169 and/or *IL6* rs13306435. Of the 125 both WTs, 94, 12, 11, and 8 patients were heterozygous-*CRIM1*, heterozygous-*IL6*, homozygous-*CRIM1*, and *IL6*-and-*CRIM1* variant groups, respectively (Table 1, Fig. 1).

**Figure 1.**
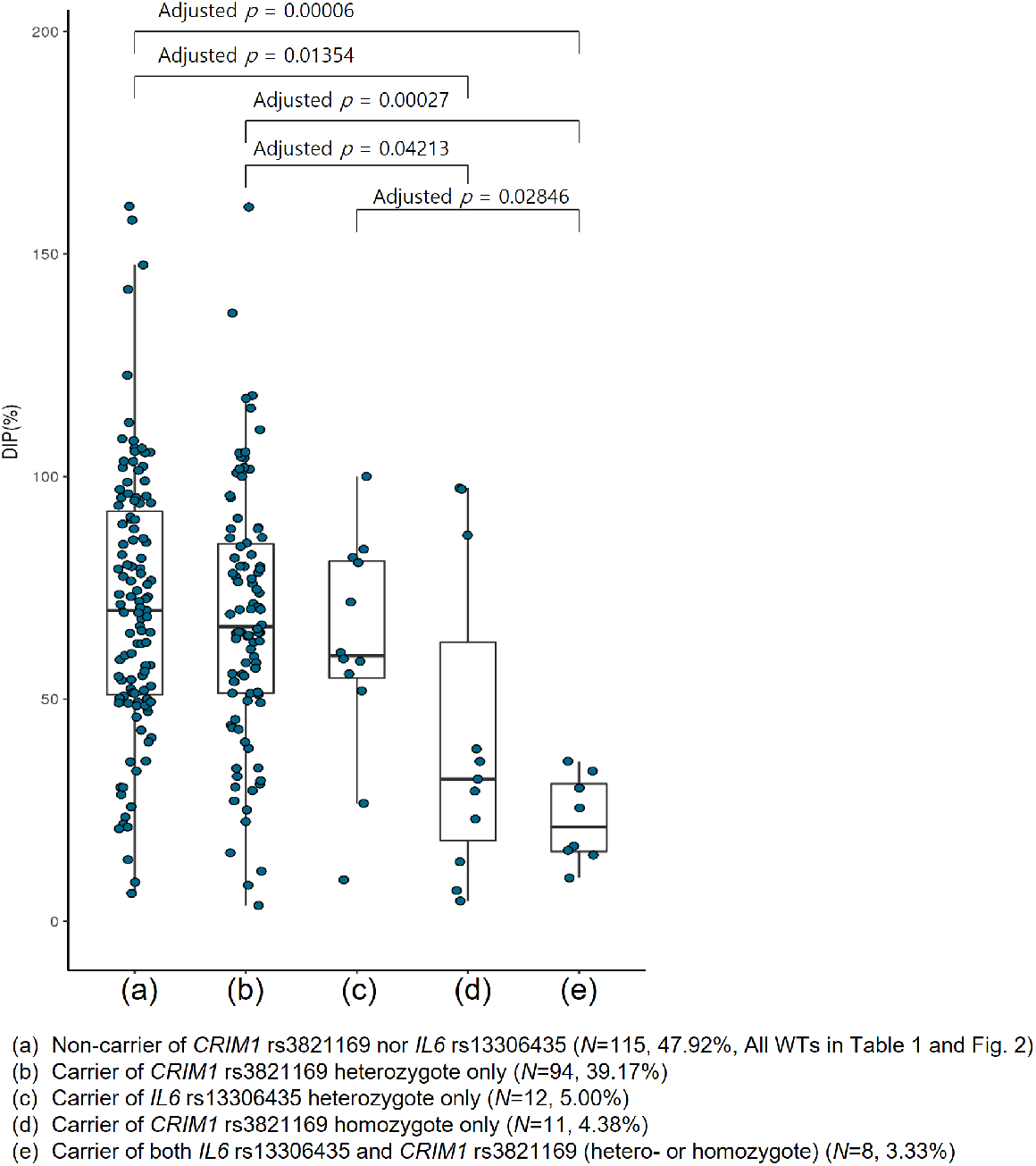
Distribution of the tolerated last-cycle 6-MP DIPs (%) of *CRIM1* rs3821169- and/or *IL6* rs13306435-variant carrier vs. non-carrier subgroups among 240 pediatric ALL subjects with both *NUDT15* and *TPMT* WTs. Of the 320 pediatric ALL patients, we excluded 80 carriers of either *NUDT15* or *TPMT* variant to obtain 240 both *NUDT15*–and-*TPMT* WT subjects. Both (a) non-carrier group of *CRIM1* rs3821169 nor *IL6* rs13306435 (*N*=115, 47.92%, i.e., ‘All WTs’ in Table 1 and Fig. 2) and (b) carrier group of *CRIM1* rs3821169 heterozygote only (*N*=94, 39.17%) showed significantly higher thiopurine tolerance than carrier groups of (d) *CRIM1* rs3821169 homozygote (*N*=11, 4.38%) (adj. *p*<0.05, posthoc Tukey) and of (e) both *IL6* rs13306435 and *CRIM1* rs3821169 (*N*=8, 3.33%) (adj. *p*<0.0005, posthoc Tukey) by one-way ANOVA (*p*=0.0001). (c) Carrier group of *IL6* rs13306435 heterozygote variant only (*N*=12, 5.00%) also showed significantly higher thiopurine intolerance than (c) that of both *IL6* and *CRIM1* hetero/homozygote (adj. *p*<0.05, posthoc Tukey). No carrier of *IL6* homozygote was found and only one subject carried both *IL6* heterozygote and *CRIM1* homozygote variants (DIP = 9.7%). Thiopurine intolerance was measured by the last-cycle 6-MP DIP(%) among the 240 pediatric ALL patients with both *NUDT15* and *TPMT* wilde type genes to control their effect on thiopurine intolerance. ALL: Acute Lymphoblastic Leukemia, WT: Wild Type, DIP (%): Dose Intensity Percentage, \**p*<0.05 and \*\**p*<0.01, post-hoc Tukey test after one-way ANOVA

We used all WTs as non-PGx controls (*N*=115) for the following analysis. We replicated previous PGx findings of *NUDT15, TPMT*, and homozygous-*CRIM1*. The tolerated 6-MP DIPs of non-WTs (i.e., IM or PM) for *NUDT15* (47.1±30.5%, *N*=72) and/or for *TPMT* (56.6±33.6%, *N*=9) were significantly lower than that of all WTs (71.3±29.6%, *N*=115) (*p*<0.001, Table 1). The homozygous-*CRIM1* group (dark blue circle in Fig. 2) exhibited significantly lower 6-MP DIP of than all WTs before (*N*=16, 44.6±35.2%) or after (*N*=11, 42.3±35.0%) controlling the five subjects with *NUDT15* (59.76±37.24%) or *IL6* (9.77%) variants.

**Figure 2.**
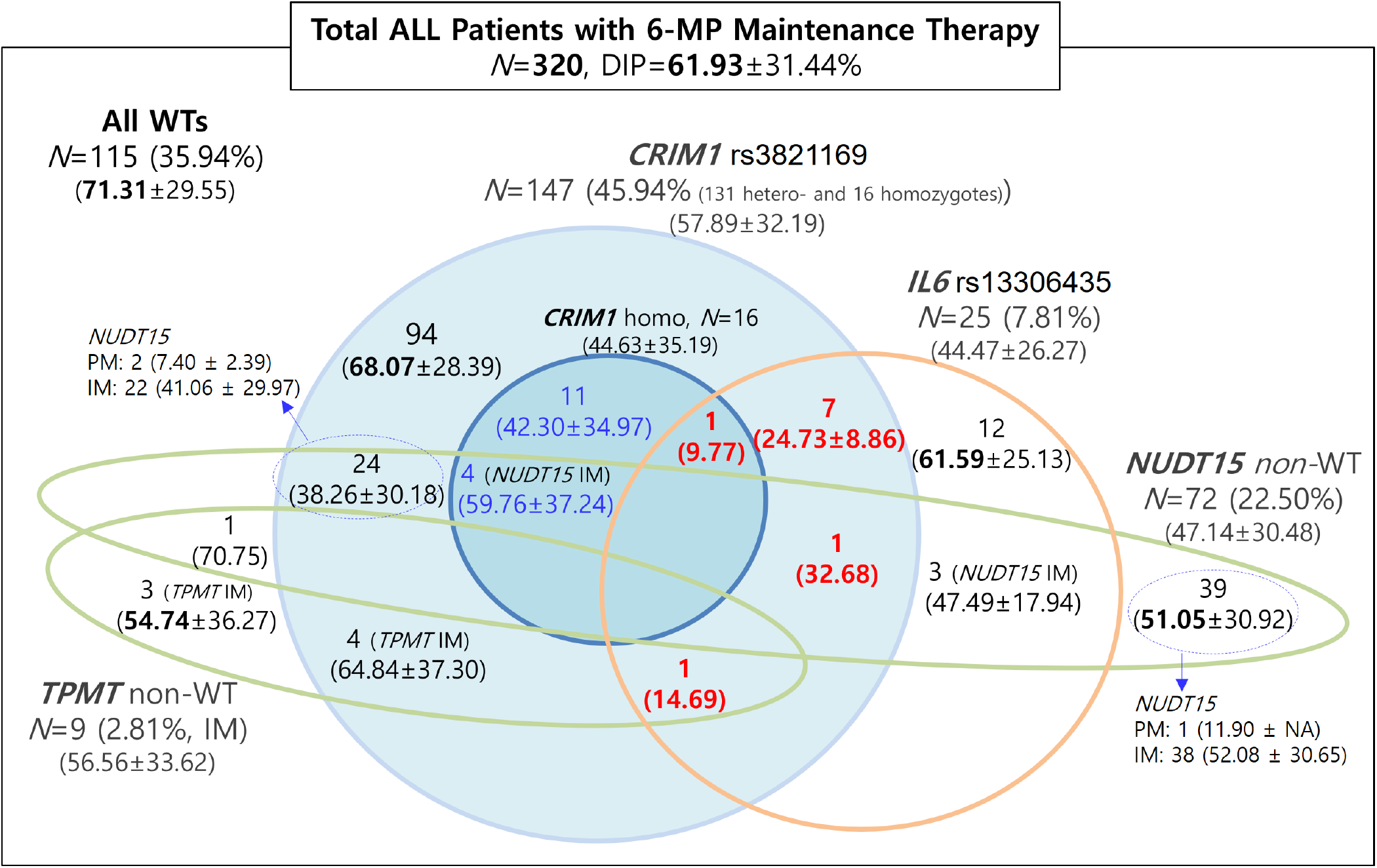
Distribution of the last-cycle 6-MP DIP of pediatric ALL patients according to *NUDT15, TPMT, CRIM1* and *IL6* pharmacogenetic subgroups (N=320). Green circles depict *NUDT15* and *TPMT* metabolizer phenotypes and blue and orange circles represent *CRIM1* rs3821169 and *L6* rs13306435 genotype subgroups, respectively. Of the 320 patients, 115 with no pharmacogenetic variants exhibited higher 6-MP DIPs (71.31%) than 72 *NUDT15* (47.14%), 9 *TPMT* (56.56%), 147 *CRIM1* (57.89%), and 25 *IL6* (DIP=44.47%) non-WTs. Subjects with both *CRIM1* and *IL6* variants (*N*=10, 3.13%) exhibited the lowest DIPs (9.77∼32.68%, in red numbers). Numbers are number of subjects and 6-MP DIPs (mean±S.D.). ALL: Acute Lymphoblastic Leukemia, WT: Wild Type, non-WT: non wild-type, i.e., poor or intermediate metabolizers for *NUDT15* and *TPMT*, DIP: Dose Intensity Percentage.

To control the PGx effect of *NUDT15, TPMT*, and homozygous-*CRIM1* on thiopurine toxicity, we extracted 228 samples who are non-carriers of these variants. We explored them for further discovery of novel PGx variant. We found that carriers of a novel variant, *IL6* rs13306435 (*N*=19, 48.0±27.3%), exhibited significantly lower 6-MP DIP than non-carriers (*N*=209, 69.9±29.0%) by Student’s *t*-test (*p*=0.0016) and multiple covariates linear regression (*p*=0.0028). Further, of the 19 carriers, we found that seven patients with both *IL6* rs13306435 and heterozygous-*CRIM1* showed significant decrease in 6-MP DIP (24.7 ± 8.9%) compared to the 12 patients harbouring *IL6* rs13306435 variant only (61.6 ± 25.1%) (brown circle in Fig. 2). Potential interplay between *IL6* and *CRIM1* variants were suggested, which was further supported by the following finding that the seven patients with both *IL6* and *CRIM1* variants showed significantly lower 6-MP DIP (24.7 ± 8.9%) than 94 heterozygous-*CRIM1* carriers (68.1±28.4%, light blue circle in Fig. 2).

### Interplay of *IL6* and *CRIM1* variants in thiopurine toxicity

Figure 1 exhibits the distributions of the last-cycle 6-MP DIPs (%) of 115 all WTs (Fig. 1(a)) and 125 *CRIM1* and/or *IL6* variant carriers who are both WTs (for *NUDT15* and *TPMT*) and consist of four PGx groups; heterozygous-*CRIM1* (*N*=94), heterozygous-*IL6* (*N*=12), homozygous-*CRIM1* (*N*=11), and *IL6*-and-*CRIM1* (*N*=8) (Fig. 1(b)-(e), see Table 1). Homozygous-*CRIM1* and *IL6*-and-*CRIM1* (44.6±35.2% and 24.7±8.9%, respectively, Fig. 1(d, e)) groups showed significantly lower 6-MP DIPs than all-WT and heterozygous-*CRIM1* groups (71.3±29.6% and 68.1±28.4%, respectively, Figs. 1(a, b)) by one-way ANOVA (*p*=0.0001; adj. *p*<0.05 posthoc Tukey). Further, *IL6*-and-*CRIM1* group showed significantly lower 6-MP DIP (44.6±35.2%) than heterozygous-*IL6* group (61.6±25.1%; adj. *p*<0.05, posthoc Tukey) (Fig. 1(c, e)). All of the 10 (=7+1+1+1) patients with both *IL6* and *CRIM1* variants (in red numbers in Fig. 2) exhibited the lowest DIPs (9.77∼32.68%) among all subgroups of the whole PGx groups. Thus, significant interplay between *IL6* and *CRIM1* in thiopurine toxicity was suggested.

Clinically more relevant is the evaluation of the magnitude of the actual decrease in 6-MP DIP (%) tolerated by patients than mere statistical significance affected by study sample size and biomarker prevalence. Table 1 demonstrates that about one quarters of homozygous-*CRIM1* (27.3%, 3/11) and *IL6*-and-*CRIM1* (25.0%, 2/8) groups were intolerant less than 25% of the planed DIP, increasing the risk of thiopurine therapeutic failure and relapse of leukemia. These intolerance levels are comparable to those of the long-known *NUDT15* (29.2% = 21/72) and *TPMT* (22.2%, 2/11) non-WTs (Table 1) published in the current CPIC guideline. Furthermore, when we raised the DIP cutoff from 25% to 35%, the quarter proportions of homozygous-*CRIM1* and *IL6*-and-*CRIM1* groups went up to 54.6% (6/11) and 87.5% (7/8), respectively, which far exceeded 38.9% and 33.3% of *NUDT15* (28/72) and *TPMT* (3/9) non-WTs, respectively. Please notice that only 6.1 (7/115) and 10.5% (12/115) of all WTs were intolerant less than 25% and 35% of the planed DIP (Table 1).

### Interethnic variabilities in carrier frequencies and molecular phenotypes

Both *NUDT15* and *TPMT* shows wide inter-ethnic variabilities. Table 2 exhibits inter-ethnic variabilities of the PGx variants and molecular phenotypes of the four thiopurine pharmacogenes computed from the 2504 subjects of the 1000 Genomes Project [9]. *NUDT15* non-WT (i.e., IM or PM) is popular in East (22.6%) and South (13.9%) Asians but rare in Europeans and Africans (<1%). In contrast, *TPMT* non-WT is popular in Europeans (8.0%) and Americans (13.3%) but relatively rare in Asians (<5.0%).

Novel PGx variant, *CRIM1* rs3821169, demonstrates remarkably high minor allele frequency (T=0.255) and carrier prevalence (43.7%, 220/504) in East Asians. Table 2 also shows that 6.5% of East Asians harbor homozygous *CRIM1* rs3821169 variant, which can hardly be found in other populations (<1.0%). In contrast, *IL6* rs13306435 is widely distributed with the highest carrier frequency of 15.0% in Americans and 3.0% among Asian and European populations. It is rare in South Asian and African populations (<1.0%). The carrier frequency of both *IL6*-and-*CRIM1* variants were 2.0% and 1.2% of East Asian and American populations, respectively.

### Single- and multi-gene prediction performances of *IL6* and *CRIM1*

We performed ROC analysis of GVB-based single- and multi-gene models for predicting the last-cycle 6-MP DIP (%) using 240 both WTs for *NUDT15* and *TPMT* to control their long-known PGx effects. Figure 3 demonstrates that (b) GVB^*CRIM1*^ outperformed (a) GVB^*IL6*^ in predicting DIPs at all cutoff levels, probably due to the higher variant frequency of *CRIM1* over *IL6* in the study population. Two-gene model GVB^*IL6,CRIM1*^ (Fig. 3(c)) consistently outperformed each of the single gene models (GVB^*IL6*^ and GVB^*CRIM1*^) at all cutoffs.

For comprehensive evaluation of all PGx interactions among *NUDT15, TPMT, IL6*, and *CRIM1*, we performed comprehensive ROC analysis using the whole 320 pediatric ALL patients (Fig. 4). Among the four single-gene model in Figure 4 (a, b, d, e) GVB^*NUDT15*^ outperformed others at all cutoffs, probably due to *NUDT15*‘s high prevalence and strong metabolic impact of on thiopurine toxicity. Two-gene models (Fig. 4 (c, f)) consistently outperformed each of the corresponding single-gene counterparts, i.e., AUCs of GVB^*NUDT15,TPMT*^ > GVB^*NUT15*^ > GVB^*TPMT*^ and of GVB^*IL6,CRIM1*^ > GVB^*CRIM1*^ > GVB^*IL6*^ at all cutoff levels. Three-gene models created by adding *IL6* or *CRIM1* to the traditional *NUDT15* and *TPMT* model also consistently improved the prediction accuracies (Fig. 4(g, h)). The final four-gene model in Figure 4(i) outperformed all other models in predicting DIPs at all cutoff levels. Moreover, it is worth noting that the ROC curves across eight DIP cutoffs in Figure 4 exhibited ‘dose-response relationships’, i.e., GVB score’s prediction power (measured by AUC) increases as a function of the severity of thiopurine intolerance (measured by DIP). For instance, the final four-gene model’s AUC increases as a function of decreasing DIP (%) (i.e., AUC^<15%^ = 0.757, AUC^<25%^ = 0.748, AUC^<35%^ = 0.711, AUC^<45%^ = 0.716, AUC^<60%^ = 0.646, and AUC^<80%^ = 0.592 in a descending order, Fig. 4(i)).

### Evaluation of clinical validity and utility of star allele and GVB method

We systematically compared the clinical utility as well as clinical validity of traditional star (*) allele-based and GVB-based methods for preventing thiopurine toxicity. Table 3 demonstrates the measures of clinical validity, i.e., sensitivity, specificity, PPV, and NPV, and of potential population impact (or clinical utility), i.e., RR, PAF, NNT, and NNG along with pharmacogenetic association (odds ratio (OR) of different prediction modes [14]. Because no designated star allele for *IL6* or *CRIM1* is available yet, star allele-based molecular phenotyping was not applicable for these novel genes. We used GVB method.

**Table 3.**
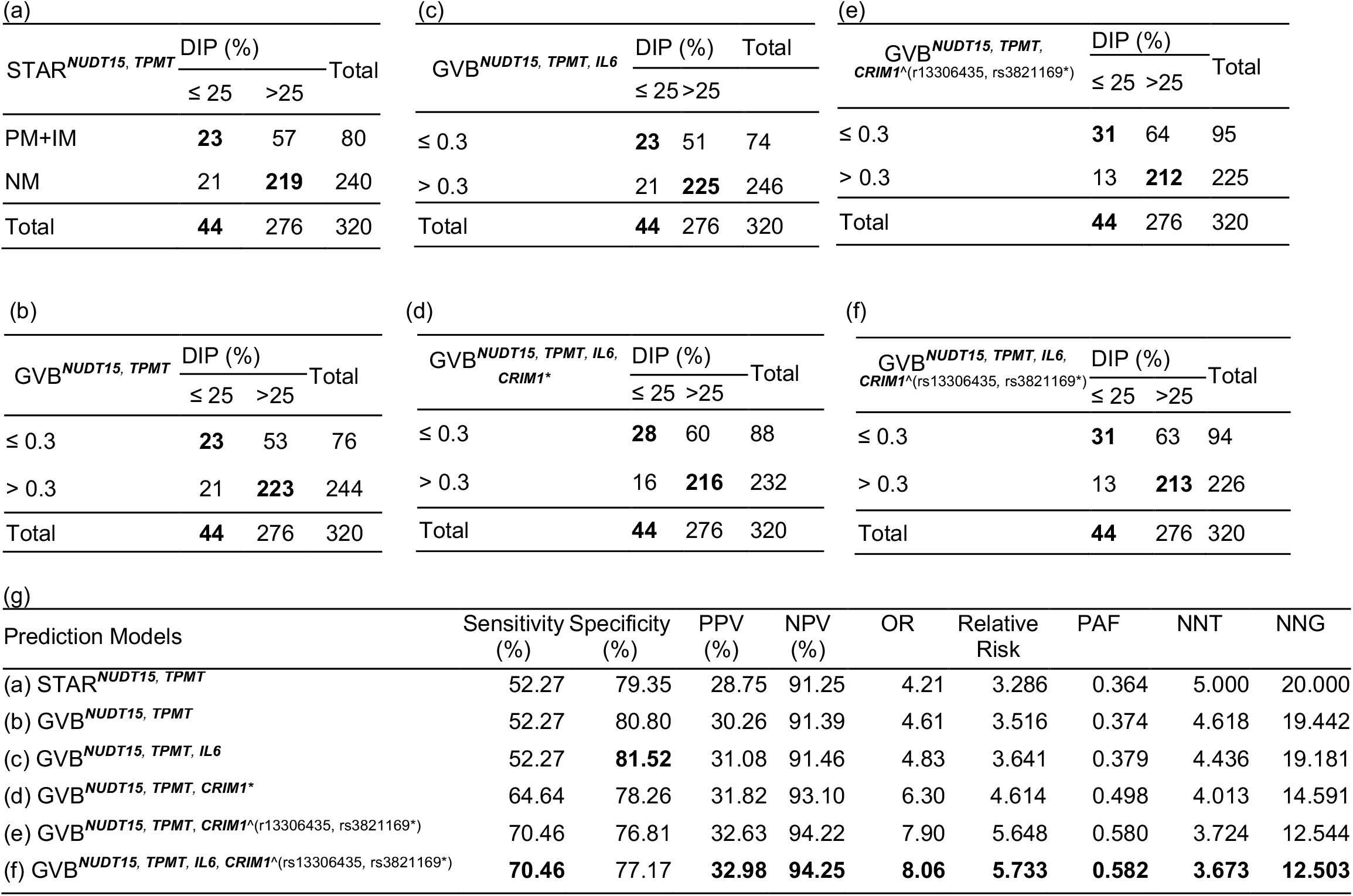
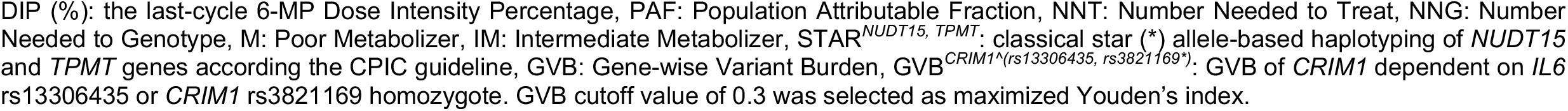
Contingency Tables for predicting thiopurine intolerance (DIP < 25%) of two-, three-, and four-gene models in pediatric ALL patients (*N*=320).

| (a) |  |  |  | (c) |  |  |  | (e) |  |  |  |  |
| --- | --- | --- | --- | --- | --- | --- | --- | --- | --- | --- | --- | --- |
| STAR<sup>NUDT15, TPMT</sup> | DIP (%) |  | Total | GVB<sup>NUDT15, TPMT, IL6</sup> | DIP (%) |  | Total | GVB<sup>NUDT15, TPMT, CRIM1<sup>Δ</sup>(r13306435, rs3821169*)</sup> | DIP (%) |  | Total |  |
|  | ≤ 25 | >25 |  |  | ≤ 25 | >25 |  |  | ≤ 25 | >25 |  |  |
| PM+IM | 23 | 57 | 80 | ≤ 0.3 | 23 | 51 | 74 | ≤ 0.3 | 31 | 64 | 95 |  |
| NM | 21 | 219 | 240 | > 0.3 | 21 | 225 | 246 | > 0.3 | 13 | 212 | 225 |  |
| Total | 44 | 276 | 320 | Total | 44 | 276 | 320 | Total | 44 | 276 | 320 |  |
| (b) |  |  |  | (d) |  |  |  | (f) |  |  |  |  |
| GVB<sup>NUDT15, TPMT</sup> | DIP (%) |  | Total | GVB<sup>NUDT15, TPMT, IL6, CRIM1*</sup> | DIP (%) |  | Total | GVB<sup>NUDT15, TPMT, IL6, CRIM1<sup>Δ</sup>(rs13306435, rs3821169*)</sup> | DIP (%) |  | Total |  |
|  | ≤ 25 | >25 |  |  | ≤ 25 | >25 |  |  | ≤ 25 | >25 |  |  |
| ≤ 0.3 | 23 | 53 | 76 | ≤ 0.3 | 28 | 60 | 88 | ≤ 0.3 | 31 | 63 | 94 |  |
| > 0.3 | 21 | 223 | 244 | > 0.3 | 16 | 216 | 232 | > 0.3 | 13 | 213 | 226 |  |
| Total | 44 | 276 | 320 | Total | 44 | 276 | 320 | Total | 44 | 276 | 320 |  |
| (g) |  |  |  |  |  |  |  |  |  |  |  |  |
| Prediction Models |  |  |  | Sensitivity (%) | Specificity (%) | PPV (%) | NPV (%) | OR | Relative Risk | PAF | NNT | NNG |
| (a) STAR<sup>NUDT15, TPMT</sup> |  |  |  | 52.27 | 79.35 | 28.75 | 91.25 | 4.21 | 3.286 | 0.364 | 5.000 | 20.000 |
| (b) GVB<sup>NUDT15, TPMT</sup> |  |  |  | 52.27 | 80.80 | 30.26 | 91.39 | 4.61 | 3.516 | 0.374 | 4.618 | 19.442 |
| (c) GVB<sup>NUDT15, TPMT, IL6</sup> |  |  |  | 52.27 | 81.52 | 31.08 | 91.46 | 4.83 | 3.641 | 0.379 | 4.436 | 19.181 |
| (d) GVB<sup>NUDT15, TPMT, CRIM1*</sup> |  |  |  | 64.64 | 78.26 | 31.82 | 93.10 | 6.30 | 4.614 | 0.498 | 4.013 | 14.591 |
| (e) GVB<sup>NUDT15, TPMT, CRIM1<sup>Δ</sup>(r13306435, rs3821169*)</sup> |  |  |  | 70.46 | 76.81 | 32.63 | 94.22 | 7.90 | 5.648 | 0.580 | 3.724 | 12.544 |
| (f) GVB<sup>NUDT15, TPMT, IL6, CRIM1<sup>Δ</sup>(rs13306435, rs3821169*)</sup> |  |  |  | 70.46 | 77.17 | 32.98 | 94.25 | 8.06 | 5.733 | 0.582 | 3.673 | 12.503 |

DIP (%): the last-cycle 6-MP Dose Intensity Percentage, PAF: Population Attributable Fraction, NNT: Number Needed to Treat, NNG: Number Needed to Genotype, M: Poor Metabolizer, IM: Intermediate Metabolizer, STAR<sup>NUDT15, TPMT</sup>: classical star (\*) allele-based haplotyping of *NUDT15* and *TPMT* genes according the CPIC guideline, GVB: Gene-wise Variant Burden, GVB<sup>CRIM1<sup>(rs13306435, rs3821169\*)</sup></sup>: GVB of *CRIM1* dependent on *IL6* rs13306435 or *CRIM1* rs3821169 homozygote. GVB cutoff value of 0.3 was selected as maximized Youden's index.

GVB^*NUDT15,TPMT*^ slightly outperformed STAR^*NUDT15,TPMT*^, the classical star (*) allele-based molecular phenotyping (Table 3(a, b)). Three-gene models (i.e., GVB^*NUDT15,TPMT,IL6*^ and GVB^*NUDT15,TPMT,CRIM1\**^ in Table 3(c) and (d)) also outperformed the two-gene models. Four-gene interplay model, GVB^*NUDT15,TPMT,IL6,CRIM1*^(*CRIM1,IL6*)^, presented the best performances for all of the eight measures of clinical validity and potential population impact (except specificity) (marked in bold numbers in Table 3(g)).

Addition of *IL6* and *CRIM1* to create the final four-gene model integrating both common and rare alleles markedly improved PAF from 0.36 to 0.58 as well as RR (3.29 to 5.73) and OR (4.21 to 8.06). PAF is the proportion of events that is attributed to the PGx risk factor or the maximum percentage of cases that can be prevented if individuals who test positive for the PGx variants receive different treatments. Of the 44 adverse events (DIP<25%), it might have prevented eight more patients from 23 to 31 than traditional star (*) allele-based method (STAR^*NUDT15,TPMT*^, Table 3(a) vs. (f)).

The number needed to genotype (NNG) is the number of patients that has to be genotyped to prevent one patient from having an adverse event. The NNG of 20 and an NNT of 5 of the traditional STAR^*NUDT15,TPMT*^ mean that for every 20 patients that are genotyped, 5 patients will learn that they test positive and need to receive alternative treatment to prevent adverse event (DIP<25%) in one. Adding *IL6* and *CRIM1* to the traditional *NUDT15* and *TPMT* testing to create GVB^*NUDT15,TPMT,IL6,CRIM1*^(*CRIM1,IL6*)^ may require only 12.5 patients (37.5% improvement of NNG) to be genotyped to return 3.7 test-positive patients (26.0% improvement of NNT) receiving alternative treatment to prevent adverse event in one (Table 2(g)).

## Discussion

Interplay between *IL6* and *CRIM1* variants on thiopurine-induced hematological toxicity was investigated in 320 pediatric ALL patients. *IL6* has been known to behavior as a neutrophil protector. There was an inverse correlation between *IL6* levels and neutrophil apoptosis (*r* = (-)0.855 and *p* < 0.007), but this was not the case for other cytokines. The antiapoptotic effect of osteomyelitis sera was reversed with anti-*IL6* antibodies and was reproduced with recombinant human *IL6* [28]. Decreased neutrophil counts have been reported in trials of tocilizumab in rheumatoid arthritis patients [29, 30]. Tocilizumab is a recombinant monoclonal Ab that binds to soluble and membrane-bound *IL-6R* (Interleukin 6 Receptor) and inhibits *IL6* signaling pathways [31, 32]. Preclinical studies suggest that the reduction in neutrophil count may result from increased margination of circulating neutrophils into the bone marrow rather than from the drug-induced neutropenia observed with myelotoxic drugs [33, 34].

*CRIM1* (Cysteine-Rich Transmembrane BMP Regulator 1) is a cell-surface transmembrane protein that resembles developmentally important proteins which are known to interact with bone morphogenetic proteins (BMPs). A role of *CRIM1* in drug resistance has been suggested by previous studies [35, 36] revealing that the level of mRNA expression of *CRIM1* is high in resistant leukemic cells. This affects the levels of BMPs, suggesting that *CRIM1* regulates the growth and differentiation of hematopoietic cells. It is suggested that the interplay of *IL6* and *CRIM1* on thiopurine-induced hematological toxicity may be a pharmacodynamic effect on adverse reaction while the long-known *NUDT15* and *TPMT* are pharmacokinetic enzymes for metabolizing thiopurines.

Seven (6.1%) of the 115 all-WT patients still suffered from thiopurine toxicity. Supplementary Table S1 lists further candidate variants determined by analyzing the all WTs (*N*=115, *p*<0.05 by one-sided Student *T*-test). Of the three carriers of *FSIP2* rs191083003, two (66.7%) exhibited DIP < 25% (8.82, 21.88, and 48.54%, *N*=3). We found one more *FSIP2* rs191083003 carrier among the homozygous-*CRIM* group, who exhibited the lowest DIP of 6.94% among the whole 320 ALL cohort. The low frequency (1.25%, 4/320) of *FSIP2* rs191083003 prohibited any conclusion but to await further elucidation. Overall, the interplay of *IL6* and *CRIM1* along with the long-known *NUDT15* and *TPMT* improved PAF from 36.4% to 58.2% by considering PGx variants only in an East Asian cohort of pediatric ALL (*N*=320). The quantitative analytic approach of the present study may be applied to other ethnic groups for further discovery and evaluation of thiopurine-toxicity pharmacogenomics.

Americans show the highest allele frequency of *IL6* rs13306435 (A=0.078) among all ethnic groups (Global A=0.020, the 1000 Genomes Project, Phase 3 [9]). This high inter-ethnic variability may partially explain why rs13306435 has not yet been discovered as a biomarker for thopurine toxicity. Current research is mostly biased towards Europeans [37]. *NUDT15* rs116855232 variant, which was recently discovered in the Korean population as a strong predictor of thopurine toxicit [3], shows the highest allele frequency in East Asians (T=0.095) among all ethnic groups (Global T=0,040). Pharmacogenes by definition, unlike pathogenic disease genes, do not have an overt phenotype unless exposed to drugs. The absence of detrimental phenotypic effect from phrmacogenes may have permitted wide inter-ethnic variability and/or diversity across different ethnic groups under various evolutionary selection pressures.

The CPIC guideline for thiopurine treatment for pediatric ALL is based on star (*) allele-based haplotypes with designated molecular phenotypes of *NUDT15* and *TPMT* [6, 7]. However, CPIC does not provide general standard rules of how to combine multi-gene interactions of the categorically classified star-alleles. Novel genes like *IL6* and *CRIM1* have no designated star alleles nor molecular phenotypes yet. Quantitative GVB method has benefits over categorical star allele-based approaches. GVB quantitates single- or multi-gene PGx burden of common, rare, and novel variants into a single score to provide a comprehensive framework for further PGx discovery and evaluation of many gene interactions. A conventional single variant-based association test of rare variants requires infeasible magnitude of sample sizes [38], but approaches that aggregate common, rare, and novel variants jointly will substantially reduce a required effective sample sizes [39]. In contrast to traditional haplotyping-based method, GVB assigns a gene-level score for each pharmacogene without using population data and hence to enable unbiased PGx method especially for under-studied subpopulations.

## Data Availability

The data that support the findings of this study are available on request from the corresponding author.

## Acknowledgements

This research was supported by a grant (16183MFDS541) from Ministry of Food and Drug Safety in 2019.

## Ethical statement

The study was approved by the AMC Review Boards, the SMC Review Boards, and the SNUH Review Boards. Informed written consents for blood sampling and analyses were obtained from all participants.

---

**Supplementary Table S1.** Candidate variants among 115 ALL patients of all wild types for *NUDT15, TPMT, IL6* and *CRIM1*.

| Variant rsID | Gene | Carrier (%) | Hetero/<br>Homozygote | Carrier DIP<br>(mean ± SD) | Non-carrier DIP<br>(mean ± SD) | SIFT | CADD | ExAC | P<sup>*</sup> |
| --- | --- | --- | --- | --- | --- | --- | --- | --- | --- |
| rs191083003 | *FSIP2* | 2.6% | 3 / 0 | 26.4 ± 20.3 | 72.5 ± 28.9 | 0.01 | 26.7 | 0.003 | 0.0260 |
| rs12587478 | *KLHL33* | 5.2% | 5 / 1 | 44.9 ± 22.4 | 72.8 ± 29.3 | 0 | 25.0 | 0.040 | 0.0136 |
| rs200982819 | *SLC15A3* | 6.1% | 6 / 1 | 50.6 ± 21.1 | 72.7 ± 29.6 | 0 | 29.7 | 0.028 | 0.0164 |
| rs67877771 | *IQCG* | 32.3% | 35 / 2 | 61.9 ± 25.4 | 75.8 ± 30.5 | 0.04 | 26.2 | 0.215 | 0.0059 |
| rs61758536 | *SPAG8* | 13.9% | 16 / 0 | 60.5 ± 22.5 | 73.1 ± 30.3 | 0 | 26.0 | 0.052 | 0.0297 |
| rs34337292 | *OR9Q2* | 26.1% | 29 / 1 | 63.1 ± 24.8 | 74.2 ± 30.7 | 0 | 25.9 | 0.068 | 0.0263 |

\* *p*-value by one-sided T-test, DIP: Dose Intensity Percentage

